# Insights into sexual partners and partnerships among gay, bisexual and other men who have sex with men in the United Kingdom: Results from the Reducing Inequalities in Sexual Health (RiiSH) behavioural survey

**DOI:** 10.64898/2025.12.05.25341714

**Authors:** Elizabeth Fearon, David Etoori, Dana Ogaz, Benjamin Weil, Will Nutland, David Reid, John Saunders, Hamish Mohammed, Catherine H. Mercer

## Abstract

**Objectives:** Sexual decision-making and STI prevention are influenced not only by individuals’ characteristics, but also those of their partners and their relationship with each other. To inform tailored STI prevention and better understand STI transmission dynamics, this study aimed to understand the characteristics of sexual partners and partnerships among UK GBMSM, for whom there has been limited information beyond partner numbers.

**Methods:** Working with community-based organisations, we co-designed a sexual partnerships module for the UK’s annual online cross-sectional community-based RiiSH GBMSM survey, undertaken November-December 2023. We collected data about ≤3 most recent male sex partners since August 2023 and analysed variations in relative demographic characteristics (age, ethnicity), sexual behaviours (sexual practices, condom use, HIV-PrEP use) and communication (talking about sexual health, sharing HIV status) by partner type.

**Results:** Altogether 1106 participants described 2342 partners: 12%, 20%, 11% and 57% reporting 0, 1, 2 and 3 partners, respectively, of varying combinations. Casual partner types were most commonly reported (43%) vs. one-time (36%), established (14%) and ‘uncertain’ relationships defined (7%). Concurrency was common with the mean number of ongoing partnerships >1 (1.23, 95%CI:1.17-1.30). Age-mixing varied by partner type, from median 2 (established partners) to 10 (one-time partners) years difference.

Sexual health communication differed by partner type. While highest among established partners, it was twice as common to be unaware of one-time partners’ HIV status and not to have discussed sexual health (both 68%) vs. among casual partners (30% and 28%, respectively). Condom use was low across all partnership types, though HIV-PrEP was used in 49% of one-time partnerships involving condomless anal intercourse.

**Discussion:** GBMSM in the UK have diverse types of male sexual partners and partnerships, which vary in their demographic mixing, sexual behaviour and sexual health communication. STI prevention and sexual wellbeing programmes targeting GBMSM should consider partnership and partner type.

**Key messages:** *What is already known:* Sexual behaviours vary across individuals and also partnership types. Understanding the timing, behaviours and characteristics of partnerships, and therefore the sexual contact network, may inform STI prevention efforts.

*What this study adds:* This study of GBMSM from a large, UK-wide community sample show their sexual partnerships are diverse in their behaviour, characteristics and communication, varying both between and within individuals.

*What are the implications:* Further research and practice should consider partnership type when designing and delivering STI prevention interventions to maximise their effectiveness among GBMSM and potentially more broadly.

## Introduction

Sexual wellbeing and the prevention of sexually transmitted infections (STIs) is influenced not only by the characteristics of individuals, but also those of their partners and the relationships that they have with each other. Surveys among many populations have found that sexual behaviours, STI prevention(1–3) and communication(4) differ across partnership types, even within the same individual. Relative characteristics between partners such as age or ethnicity, can also influence sexual health decision-making(5).

Partnerships link together into sexual networks, the structure of which influences STI transmission patterns, while individual position within the network influences risk of acquiring STIs(6, 7). While some network characteristics, such as the cumulative distribution of numbers of partners (‘degree distribution’), can be measured by asking individuals about themselves, other important characteristics, such as homophily – the extent to which people form partnerships with others like or unlike themselves - require data about the characteristics of specific partners. If populations with systematically under-resourced access to sexual healthcare are more likely to form within-group partnerships, effects of these inequalities could compound(8).

Partnership concurrency – whereby partnerships overlap with each other in time - has been found to increase STI epidemic growth(9), but its influence depends also on partnership duration and length of overlap and both are difficult to measure in the absence of specific reported partnership data.

GBMSM globally are disproportionately affected by high STI incidence(10) including in the UK(11). New approaches to bacterial STI prevention are becoming available (including DoxyPEP(12), taken on a per event basis), but understanding their potential population impact and supporting effective use requires understanding of the types of partnerships that GBMSM are having.

There is little quantitative information about the characteristics of recent sexual partners and partnerships among GBMSM in the UK. Britain’s population-representative Natsal surveys, which do collect data about recent sexual partners(13), do not have sufficient sample size to explore in detail GBMSM partnerships (n=148 men reporting at least one same sexual partner in the previous year in Natsal-3(14, 15)).

This study aimed to provide a first quantitative analysis of recent sexual partnerships among GBMSM in the UK. With community partners, we co-developed a new module for an online community sexual health survey to describe 1) recent male partners/partnerships by type of partnerships and timing; 2) measures of demographic similarity between participants and their reported partners; and 3) sexual behaviours and communication across different types of partner.

## Methods

### Study design

This study used data collected in the seventh round of the online community-based Reducing inequalities in Sexual Health (RiiSH) survey (see (16)), conducted November 7- December 6, 2023.

### Setting and sampling

GBMSM were recruited through community partners, social networking sites (Facebook, Twitter, Instagram) and geospatial dating applications (Grindr). Eligibility included UK resident, ≥16 years, men (cisgender/transgender) or gender diverse individuals assigned male at birth (AMAB) who reported sex with a man (cisgender/transgender) or gender diverse individual AMAB in the past year. Online consent was obtained from all participants. No financial incentives were offered. The survey was administered using Snap Surveys platform (www.snapsurveys.com).

### Data collection

Participants self-reported demographic characteristics (age, gender, ethnicity, education, employment), STI/HIV testing, PrEP use, sexual health service use, sexual relationships and behaviours, use of chemsex drugs, new STI interventions and sexual partners and partnerships.

We collaborated between academic, public health agency and a community-based organisation (CBO) to co-design a new partnership module. We held one face-to-face community member workshop and two online community-based organisation workshops (8 CBOs) during summer 2023 to understand: types/typologies of sexual partners; how partners are met; partner question topics that were/were not acceptable and/or feasible to answer; and topic priorities.

A community-generated mapping of partnership types was simplified for survey implementation to 1) established partners (e.g. boyfriend, husband, partner); 2) casual partners (e.g. friend-with-benefits, fuck buddies); 3) one-time partners and 4) uncertain partners, for whom categorization was not yet clear. Where not generated via community engagement, we adapted existing questions from surveys such as Natsal(14).

After reporting total numbers of male/female sexual partners since August 2023 (previous three months), participants reported information about up to three (if applicable) most recent male sexual partners within this same period: partnership type, dates of first/last sex, whether sex was expected again in future (partnership was likely ‘ongoing’) and whether paid sex occurred in the partnership. Sex was defined as any physical genital contact. Participants were then asked additional questions about their most recent partner, including age, ethnicity, residence, how met, sexual behaviours, condom and HIV PrEP use (as applicable), communication, and HIV status sharing. For those reporting >1 partner since August 2023, additional questions for the second and third most recent partner, as applicable, were also optionally answered.

### Ethics

The study received ethical approval from the UKHSA Research and Ethics Governance Group (REGG; ref: R&D 524).

### Data analysis

Descriptive analyses of demographic, behavioural, and partnership characteristics were calculated by partnership type. Momentary degree, defined as the number of partnerships of duration (up to 3) reported on the day of interview, and Poisson regression used to estimate mean and 95% confidence intervals. Partnership age was calculated for recently initiated partnerships (within the previous 5 years) as either the interval between first sex and interview date (if participant expected sex again in the future) or the interval between first and most recent date of sex (sex not expected again in the future). Partnership overlaps were defined as any overlapping time-spans of partnerships (first to most recent sex date or survey date if ongoing), including one-time partners, since August 2023. We examined the participant-partner age difference distribution and estimated mean age difference by partner type using linear models. We estimated ethnicity homophily by taking the ratio of reported number of same-ethnicity partnerships compared to the ‘expected’ number assuming the population of potentially available partners matched the ethnicity distribution of all partners reported in the survey, stratified by participant ethnicity.

Analysis was conducted using Stata 18.

## Results

There were 1106 eligible participants, half recruited via Grindr advertising, 23% from Instagram, 19% from Facebook and 7% from community-cascaded links. Altogether 138 (12%) reported no physical sex since August 2023. There were 968 participants reporting detailed information about 1 sexual partner (100% of those eligible), 746 participants reporting sex with 2 or more partners of whom 222 (70%) reported optional detailed information about both partners and 632 participants who reported 3 or more partners of whom 391 (62%) reported optional detailed questions about all three (Appendix Figure A1). This totaled n=2342 partners about whom partnership type, dates of sex and whether paid sex occurred were recorded and 1879 partners about whom additional detailed questions were answered. These represent 24-33% (19-25% in detail) of the 7400-9800 total partnerships reported to have occurred since August 2023 (Appendix Table A1).

### Participants’ characteristics

Participants had a median age of 44 years (IQR: 34, 54), were predominantly of White ethnicity (984, 89.0%), born in the UK (860, 77.8%), highly educated (691, 62.5% with degree), and employed full-time (713, 64.5%). 143 (12.9%) participants reported living with HIV, and 530 (47.9%) had ever used HIV-PrEP.

### Numbers of sexual partners

In the previous 3 months, 226 (20.4%) participants reported having one male partner (including cis and trans men), 110 (9.9%) 2-5 partners, 118 (10.6%) 6-9 partners, 242 (21.8%) 10-49 partners and 28 (2.5%) 50+ partners. Cumulatively, 632 (57.1%) reported at least three physical male sex partners (See Appendix Table A2). 90 participants (8.1%) reported sex with 1+ women (including cis and trans women) during this time, of whom 72 (6.5%) also reported sex with 1+ men.

### Partnership characteristics and most recent partner combinations

Of 2342 reported male sexual partnerships, 1012 (43.2%) were casual, 845 (36.1%) one-time, 326 (13.9%) established, and 159 (6.8%) uncertain (Table 1). There were 69/2342 (3%) partnerships that included paid sex (reported by 51/1106, 5% of participants).

**Table 1:**
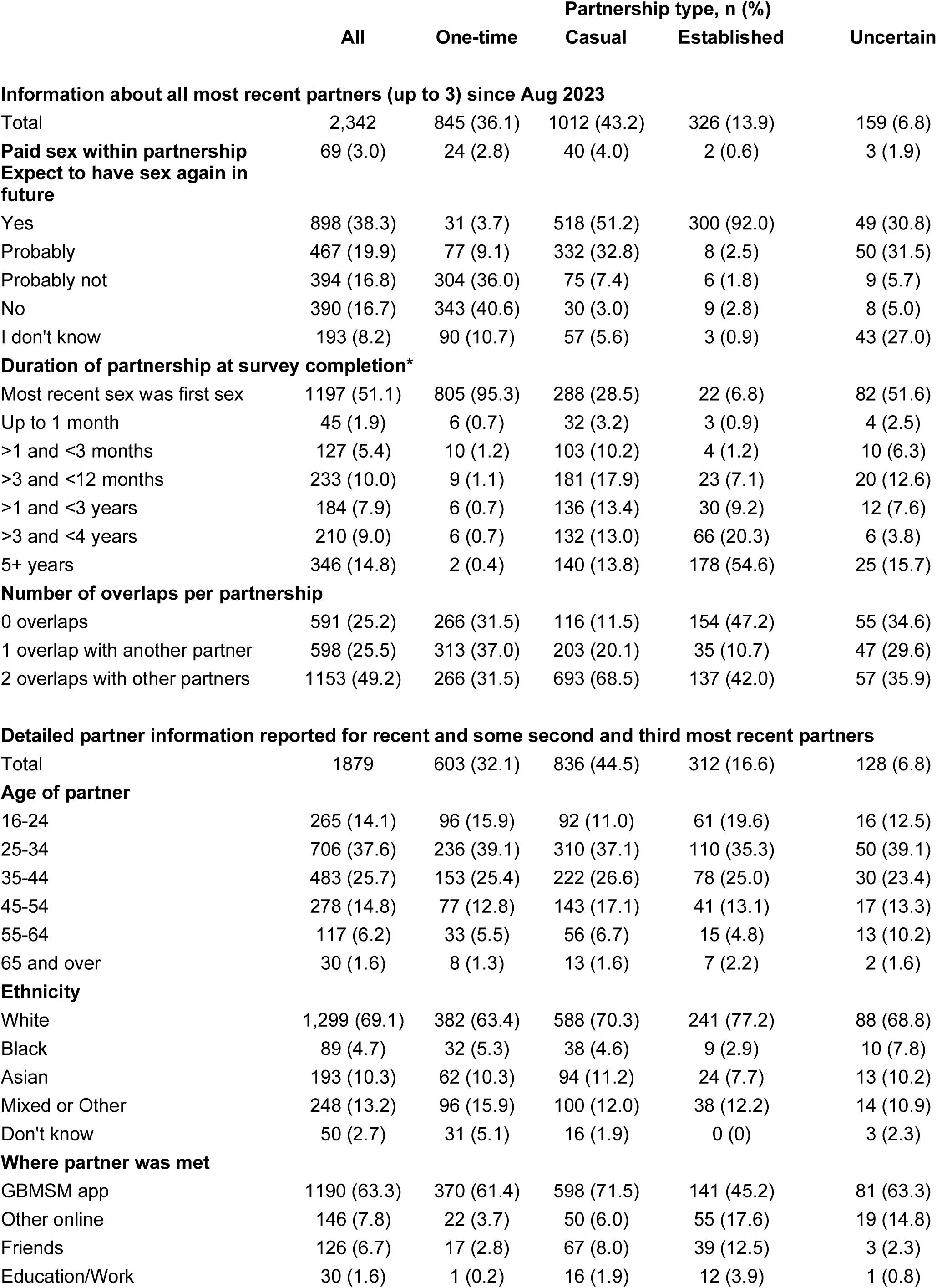
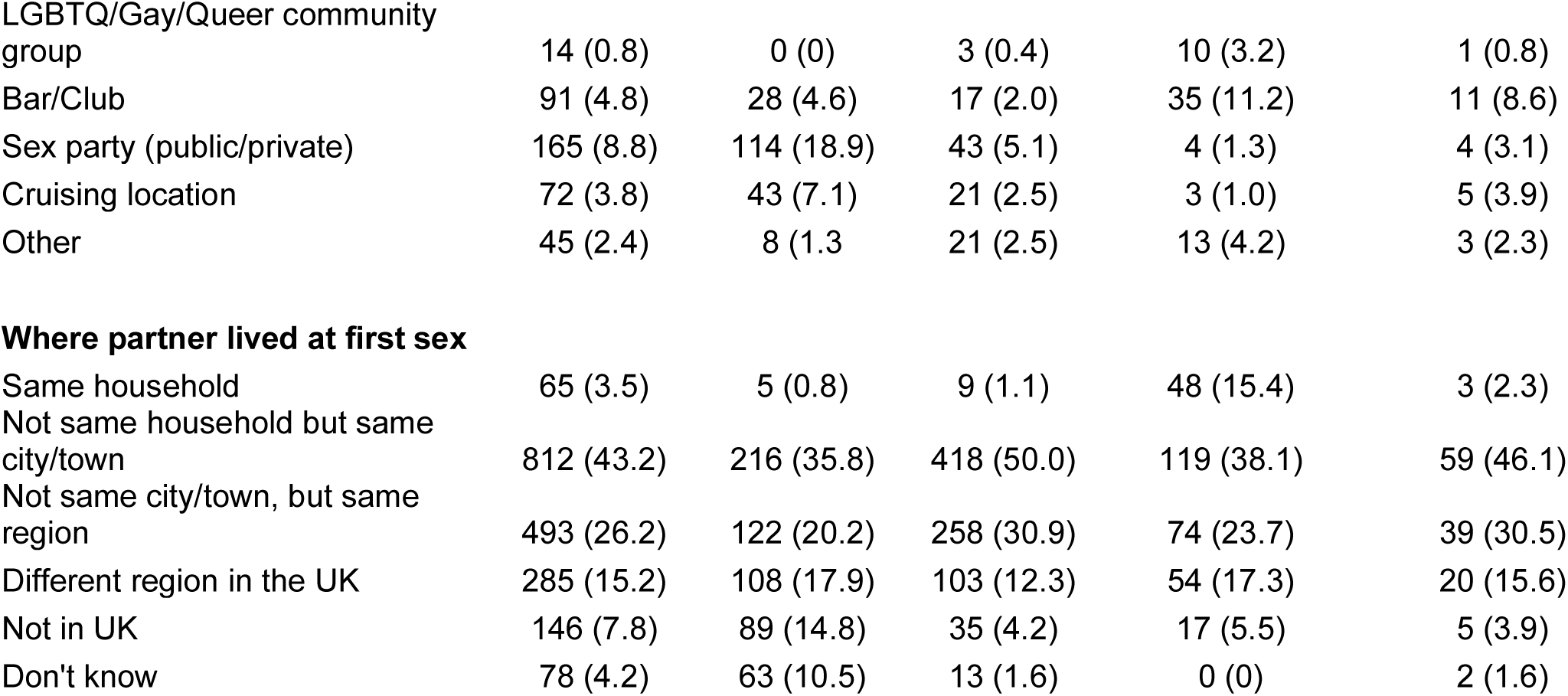
Male sexual partner and partnership characteristics reported by 968 RiiSH survey participants who reported physical sex in the previous 3 months.

|  | Partnership type, n (%) |  |  |  |  |
| --- | --- | --- | --- | --- | --- |
|  | All | One-time | Casual | Established | Uncertain |
| **Information about all most recent partners (up to 3) since Aug 2023** |  |  |  |  |  |
| Total | 2,342 | 845 (36.1) | 1012 (43.2) | 326 (13.9) | 159 (6.8) |
| **Paid sex within partnership** | 69 (3.0) | 24 (2.8) | 40 (4.0) | 2 (0.6) | 3 (1.9) |
| **Expect to have sex again in future** |  |  |  |  |  |
| Yes | 898 (38.3) | 31 (3.7) | 518 (51.2) | 300 (92.0) | 49 (30.8) |
| Probably | 467 (19.9) | 77 (9.1) | 332 (32.8) | 8 (2.5) | 50 (31.5) |
| Probably not | 394 (16.8) | 304 (36.0) | 75 (7.4) | 6 (1.8) | 9 (5.7) |
| No | 390 (16.7) | 343 (40.6) | 30 (3.0) | 9 (2.8) | 8 (5.0) |
| I don't know | 193 (8.2) | 90 (10.7) | 57 (5.6) | 3 (0.9) | 43 (27.0) |
| **Duration of partnership at survey completion\*** |  |  |  |  |  |
| Most recent sex was first sex | 1197 (51.1) | 805 (95.3) | 288 (28.5) | 22 (6.8) | 82 (51.6) |
| Up to 1 month | 45 (1.9) | 6 (0.7) | 32 (3.2) | 3 (0.9) | 4 (2.5) |
| >1 and <3 months | 127 (5.4) | 10 (1.2) | 103 (10.2) | 4 (1.2) | 10 (6.3) |
| >3 and <12 months | 233 (10.0) | 9 (1.1) | 181 (17.9) | 23 (7.1) | 20 (12.6) |
| >1 and <3 years | 184 (7.9) | 6 (0.7) | 136 (13.4) | 30 (9.2) | 12 (7.6) |
| >3 and <4 years | 210 (9.0) | 6 (0.7) | 132 (13.0) | 66 (20.3) | 6 (3.8) |
| 5+ years | 346 (14.8) | 2 (0.4) | 140 (13.8) | 178 (54.6) | 25 (15.7) |
| **Number of overlaps per partnership** |  |  |  |  |  |
| 0 overlaps | 591 (25.2) | 266 (31.5) | 116 (11.5) | 154 (47.2) | 55 (34.6) |
| 1 overlap with another partner | 598 (25.5) | 313 (37.0) | 203 (20.1) | 35 (10.7) | 47 (29.6) |
| 2 overlaps with other partners | 1153 (49.2) | 266 (31.5) | 693 (68.5) | 137 (42.0) | 57 (35.9) |
| **Detailed partner information reported for recent and some second and third most recent partners** |  |  |  |  |  |
| Total | 1879 | 603 (32.1) | 836 (44.5) | 312 (16.6) | 128 (6.8) |
| **Age of partner** |  |  |  |  |  |
| 16-24 | 265 (14.1) | 96 (15.9) | 92 (11.0) | 61 (19.6) | 16 (12.5) |
| 25-34 | 706 (37.6) | 236 (39.1) | 310 (37.1) | 110 (35.3) | 50 (39.1) |
| 35-44 | 483 (25.7) | 153 (25.4) | 222 (26.6) | 78 (25.0) | 30 (23.4) |
| 45-54 | 278 (14.8) | 77 (12.8) | 143 (17.1) | 41 (13.1) | 17 (13.3) |
| 55-64 | 117 (6.2) | 33 (5.5) | 56 (6.7) | 15 (4.8) | 13 (10.2) |
| 65 and over | 30 (1.6) | 8 (1.3) | 13 (1.6) | 7 (2.2) | 2 (1.6) |
| **Ethnicity** |  |  |  |  |  |
| White | 1,299 (69.1) | 382 (63.4) | 588 (70.3) | 241 (77.2) | 88 (68.8) |
| Black | 89 (4.7) | 32 (5.3) | 38 (4.6) | 9 (2.9) | 10 (7.8) |
| Asian | 193 (10.3) | 62 (10.3) | 94 (11.2) | 24 (7.7) | 13 (10.2) |
| Mixed or Other | 248 (13.2) | 96 (15.9) | 100 (12.0) | 38 (12.2) | 14 (10.9) |
| Don't know | 50 (2.7) | 31 (5.1) | 16 (1.9) | 0 (0) | 3 (2.3) |
| **Where partner was met** |  |  |  |  |  |
| GBMSM app | 1190 (63.3) | 370 (61.4) | 598 (71.5) | 141 (45.2) | 81 (63.3) |
| Other online | 146 (7.8) | 22 (3.7) | 50 (6.0) | 55 (17.6) | 19 (14.8) |
| Friends | 126 (6.7) | 17 (2.8) | 67 (8.0) | 39 (12.5) | 3 (2.3) |
| Education/Work | 30 (1.6) | 1 (0.2) | 16 (1.9) | 12 (3.9) | 1 (0.8) |
| LGBTQ/Gay/Queer community group | 14 (0.8) | 0 (0) | 3 (0.4) | 10 (3.2) | 1 (0.8) |
| Bar/Club | 91 (4.8) | 28 (4.6) | 17 (2.0) | 35 (11.2) | 11 (8.6) |
| Sex party (public/private) | 165 (8.8) | 114 (18.9) | 43 (5.1) | 4 (1.3) | 4 (3.1) |
| Cruising location | 72 (3.8) | 43 (7.1) | 21 (2.5) | 3 (1.0) | 5 (3.9) |
| Other | 45 (2.4) | 8 (1.3) | 21 (2.5) | 13 (4.2) | 3 (2.3) |
| **Where partner lived at first sex** |  |  |  |  |  |
| Same household | 65 (3.5) | 5 (0.8) | 9 (1.1) | 48 (15.4) | 3 (2.3) |
| Not same household but same city/town | 812 (43.2) | 216 (35.8) | 418 (50.0) | 119 (38.1) | 59 (46.1) |
| Not same city/town, but same region | 493 (26.2) | 122 (20.2) | 258 (30.9) | 74 (23.7) | 39 (30.5) |
| Different region in the UK | 285 (15.2) | 108 (17.9) | 103 (12.3) | 54 (17.3) | 20 (15.6) |
| Not in UK | 146 (7.8) | 89 (14.8) | 35 (4.2) | 17 (5.5) | 5 (3.9) |
| Don't know | 78 (4.2) | 63 (10.5) | 13 (1.6) | 0 (0) | 2 (1.6) |

Among all partners, 94.5% of established partners were ‘ongoing’ (‘yes’ or ‘probably’ will have sex again in future) as were 84.0% of casual partners and 62.3% of uncertain partners. There were 12.8% of one-time partners who were classed as ‘ongoing’ and 4.2% of ‘casual’ partners with whom participants had only had sex once and did not think they would have sex with again in future, so it is possible that the same circumstances were reported differently in some cases.

Most frequent combinations of recent partner types included casual and one-time partners among those with 2 or 3+ partners in the lookback period, and established among those reporting one partner only (Appendix Figure A2).

### Partner demographics and how partners met

The majority of partners were reported to be of White ethnicity (69%), lower than the percentage of participants self-reporting as White (89%). The most common age group of partners was 25-34 years (37.6% of partners), younger than the mean participant age (44 years), Table 1.

Partners were most commonly met via GBMSM-oriented mobile apps (63.3%), with some variation by partner type. Partners of participants recruited via Grindr were more likely to have been met on a GBMSM app than those recruited via other means (72%, compared to 53% and 55% among participants recruited via Facebook or Instagram). It was not uncommon for partners to have lived in a different town or region to the participant at the time they first had sex; among one-time partners, 14.5% were living in a different country.

### Partnership durations at survey completion

Approximately half of all reported sexual partners were first-time sexual partners (51.1%). More than half of established partnerships were more than 5 years old (54.6%) and, while newer on average, more than half of casual partnerships (58.1%) were at least 3 months old.

### Sexual behaviours within partnerships

Altogether, 74.8% of partnerships had ever involved anal intercourse, highest among established partners (93.3%), Table 2. There were 330/1879 (18%) that had ever involved group sex, again highest among established partners (32.7%), though these also had the longest average duration over which sexual behaviours could accrue.

**Table 2:** Sexual behaviours and communication within partnerships.

| Types of sex ever had in relationship | Partnership type, n (%) |  |  |  |  | Percentage of variance explained by partner characteristics (95% CI)* |
| --- | --- | --- | --- | --- | --- | --- |
|  | All | One-time | Casual | Established | Uncertain |  |
| Receptive anal sex | 935 (49.8) | 209 (34.7) | 441 (52.8) | 229 (73.4) | 56 (43.8) | 56 (49-64) |
| Insertive anal sex | 872 (46.4) | 210 (34.8) | 365 (43.7) | 242 (77.6) | 55 (43.0) | 63 (55-70) |
| Anal sex (either role) | 1405 (74.8) | 375 (62.2) | 651 (77.9) | 291 (93.3) | 88 (68.8) | 66 (59-73) |
| Oral sex | 1,506 (80.2) | 447 (74.1) | 677 (81.0) | 280 (89.7) | 102 (79.7) | 55 (48-63) |
| Group sex | 330 (17.6) | 83 (13.8) | 136 (16.3) | 102 (32.7) | 9 (7.0) | 37 (29-46) |
| Mutual masturbation | 1,106 (58.9) | 287 (47.6) | 472 (56.5) | 268 (85.9) | 79 (61.7) | 54 (47-61) |
| Other penetration | 203 (10.8) | 25 (4.2) | 96 (11.5) | 73 (23.4) | 9 (7.0) | 44 (22-49) |
| Use of sex toys | 352 (18.7) | 28 (4.6) | 162 (19.4) | 153 (49.0) | 9 (7.0) | 52 (41-62) |
| **Frequency of sex since Aug 23 (among non-first-time partners, n=955)** |  |  |  |  |  |  |
| Daily | 4 (0.4) | - | 0 (0) | 4 (1.4) | 0 (0) | 78 (60-90)** |
| A few times per week | 112 (11.7) | - | 26 (4.3) | 83 (28.4) | 3 (5.2) |  |
| Once per week | 115 (12.0) | - | 43 (7.1) | 65 (22.3) | 7 (12.1) |  |
| < Once per week, > once per month | 289 (30.3) | - | 196 (32.4) | 82 (28.1) | 11 (19.0) |  |
| < Once per month | 306 (32.0) | - | 245 (40.5) | 38 (13.0) | 23 (39.7) |  |
| Varied | 129 (13.5) | - | 95 (15.7) | 20 (6.9) | 14 (24.1) |  |
| **Condomless anal sex since Aug 2023** |  |  |  |  |  |  |
| Always used a condom | 139 (7.4) | 48 (8.0) | 78 (9.3) | 8 (2.6) | 5 (3.9) | 79 (58-91)** |
| Never used a condom | 994 (52.9) | 293 (48.6) | 438 (52.4) | 196 (62.9) | 67 (52.3) |  |
| Sometimes used a condom | 131 (7.0) | 14 (2.3) | 78 (9.3) | 34 (10.9) | 5 (3.9) |  |
| No anal sex since Aug 2023 | 615 (32.7) | 248 (41.1) | 242 (29.0) | 74 (23.7) | 51 (39.8) |  |
| **Participant HIV PrEP use during partnership (among participants not LHIV, partnership included CAS), n=1491** |  |  |  |  |  |  |
| Yes, on all occasions | 661 (44.3) | 228 (49.2) | 327 (50.0) | 63 (23.3) | 43 (42.0) | 22 (15-30)** |
| Yes, on some occasions | 65 (4.4) | 15 (3.2) | 27 (4.1) | 20 (7.4) | 3 (2.9) |  |
| No | 144 (9.7) | 36 (7.8) | 45 (6.9) | 50 (18.5) | 13 (12.6) |  |
| Don't remember | 5 (0.3) | 2 (0.4) | 3 (0.5) | 0 | 0 |  |
| Never used PrEP | 616 (41.3) | 183 (39.4) | 252 (38.5) | 137 (50.7) | 44 (42.7) |  |
| **Talking to partner about sexual health** |  |  |  |  |  |  |
| Easy | 971 (51.7) | 168 (27.9) | 520 (62.2) | 219 (70.2) | 64 (50.0) |  |
| Difficult | 82 (4.4) | 19 (3.2) | 31 (3.7) | 25 (8.01) | 7 (5.5) | 45 (37-53)** |
| Have not discussed | 686 (36.5) | 396 (65.7) | 227 (27.2) | 19 (6.1) | 44 (34.4) |  |
| Variable/depends on topic | 140 (7.5) | 20 (3.3) | 58 (6.9) | 49 (15.7) | 13 (10.2) |  |
| **Knowledge of partner HIV status** |  |  |  |  |  |  |
| Same as participant | 1,014 (54.0) | 186 (30.8) | 494 (59.1) | 259 (83.0) | 75 (58.6) |  |
| Different to participant | 152 (8.1) | 14 (2.3) | 91 (10.9) | 35 (11.2) | 12 (9.4) | 38 (31-46)** |
| Don't know | 713 (37.9) | 403 (66.8) | 251 (30.0) | 18 (5.8) | 41 (32.0) |  |

\*Calculated as 100- (Intraclass Correlation Coefficient (ICC) x 100) for participant ID in an empty two-level logistic regression random intercept model.
\*\* Variables made binary for variance decomposition (At least once per month or Less than once per month; Always use condoms or Any other response; Easy to talk about sex or Any other response; Know partner relative HIV status or Do not know partner relative HIV status; Use PrEP on all occasions or Do not use PrEP on all occasions).

Among one-time partners, 62.2% of partnerships involved anal intercourse, 74.1% oral sex, 47.6% mutual masturbation and 13.8% group sex. Among these one-time partnerships, there was negative correlation between insertive and receptive anal sex (−0.211 Pearson correlation coefficient) and positive correlation between mutual masturbation and oral sex (0.32) and use of sex toys and non-penile penetration (0.31), (Appendix Figure A3).

Condoms were reported as never used during anal sex for the majority of partnerships since August 2023 (78.6%). Most (82.5%) one-time partnerships involving anal sex never included condom use.

Among participants reporting a negative most recent HIV test, HIV-PrEP was always used during 44.3% of partnerships, highest among one-time (49.2%) and casual partnerships (50.0%). HIV PrEP was used in 49% of one-time partnerships involving condomless anal intercourse.

There was substantial variation in sexual behaviour between partners even within the same participant (‘within-participant variance’, versus ‘between-participant’ variance). The percentage of variance in sexual behaviours within-participants ranged from 22% (95% CI 15-30%) in PrEP use among participants not LHIV to 79% (95% CI 58-91%) in condom use. This indicated that condom use varied relatively more among individual partners of the same participant compared to PrEP use. Different types of sex showed roughly 37% (group sex) to 66% (anal sex) variation at the within-individual level.

### Sexual health communication and HIV status disclosure

Participants reported that it was ‘easy’ to discuss sexual health with around half (51.7%) of their partners, though for around one third (36.5%) sexual health had not been discussed at all. Non-discussion was almost two-thirds (65.7%) in one-time partnerships, Table 2.

While participants reported that they were aware of the majority of their sexual partners’ HIV statuses, this awareness varied by partner type, with 33.0% awareness for one-time, 70% for casual and 94.2% for established partners.

### Concurrency in partnerships

Roughly half of all partnerships overlapped twice in time with other of the most recent 3 partners from the previous 3 months (49.2%), Table 1. One quarter overlapped with one other partner (25.5%) and one quarter not at all (25.2%). Overlaps were most common among casual partners (68.5% overlapped with two other partners), and least among established partners, 47.2% of whom were the sole male sexual partner during this time period. 68.5% of one-time partnerships occurred during the time period of another partner (including on the same day as other one-time partners). At the participant level, nearly two thirds (64.1%) reported at least one partnership time overlap among their three most recent partners.

It was common for participants to report more than one ongoing sexual partnership. At date of interview, participants reported a mean of 1.23 (95% CI: 1.17, 1.30) ongoing partnerships (partners with whom they expected sex in future) at the time of survey completion, with highest contribution from casual partnerships.

### Age differences between sexual partners

Among 2090 recent partnerships (formed 2018 onwards), there was wide variation in age differences reported between partners, (Figure 1) with participants a mean 8.5 years older than partners (IQR -19 to 1, partner relative to participant age), but with lesser differences between established partners (mean of -3.8 years, IQR -10 to 3) compared to one-time partners (mean -10.2 years, IQR -21 to 0). Younger participants more commonly reported older sexual partners, and vice versa (Appendix Figure A4).

**Figure 1:**
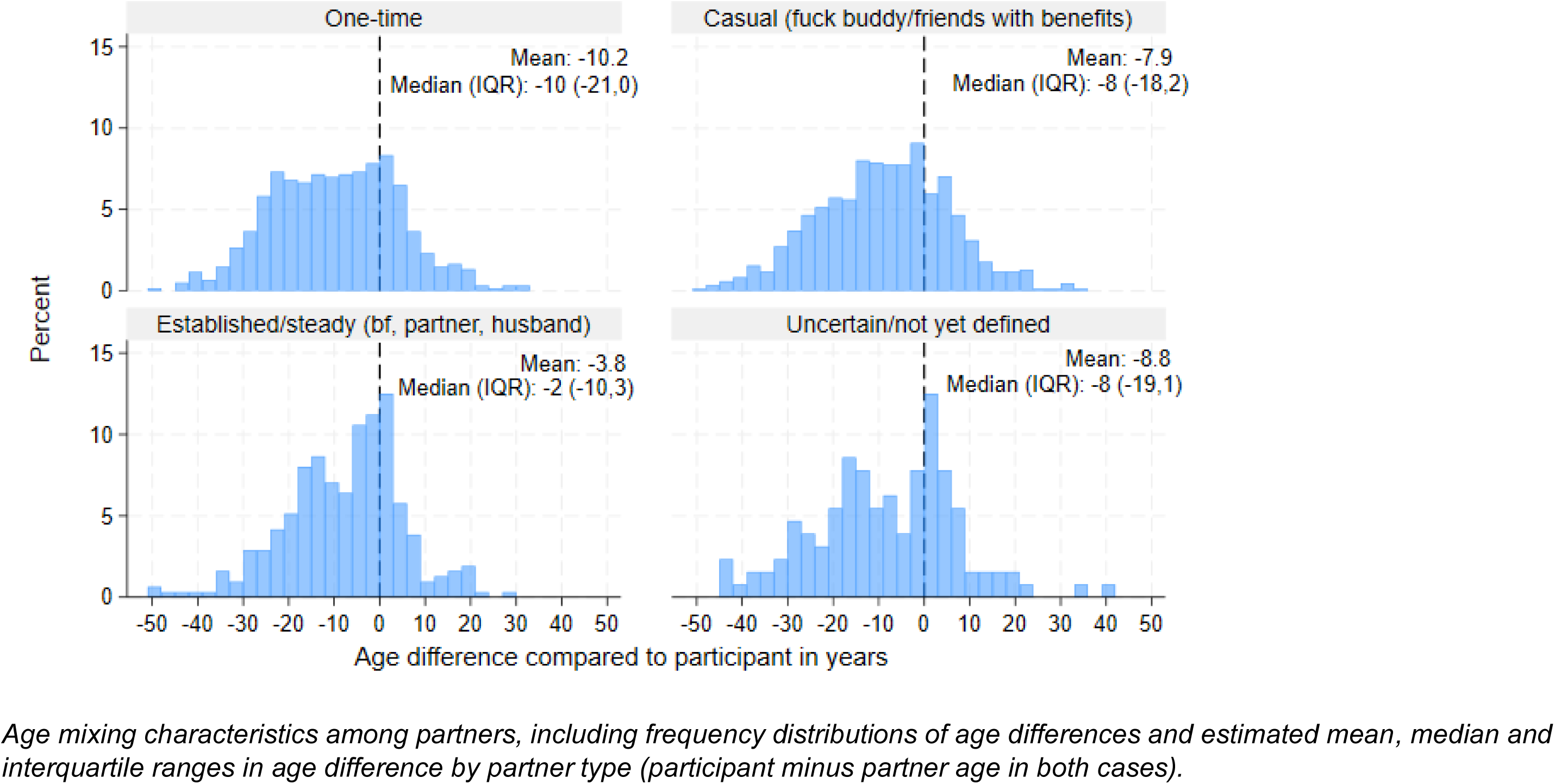
Frequency distributions of age difference in years between participant and most recent up to 3 sexual partners since August 2023, n=2090 recent partners (formed 2018 onwards) *Age mixing characteristics among partners, including frequency distributions of age differences and estimated mean, median and interquartile ranges in age difference by partner type (participant minus partner age in both cases)*.

### Ethnicity homophily among sexual partners

Participants reported that they were of the same ethnicity as their partner in 1229/1879 (65%) partnerships, Table 3. While a higher proportion of partnerships among White participants were reported to be ethnically homophilous (70%) compared to partnerships among Black, Asian and Mixed or Other ethnicity participants, these figures also reflect the ethnicity distribution among available partners in the population. Assuming that the population of available partners has the same ethnicity distribution as all reported partnerships in the survey, the ratio of observed compared to expected within-ethnicity partnerships was above 1 for Black participants (5.9, among 28 partnerships), and Asian participants (2.6, among 85 partnerships).

**Table 3:** Homophily by ethnicity in reported partnerships, n=1879 partners and n=968 participants.

| **Ethnicity of participants** | **Ethnicity of participants reporting 1+ partners, n (%)** | **Number (%) of partners reported by participants** | **Reported number of same-ethnicity partnerships R\*, n (%)** | **Ethnicity distribution of all reported partners** | **Expected number of same-ethnicity partnerships if no homophily\*\*, E** | **Ratio or reported compared to expected, R/E (95% CI)** |
| --- | --- | --- | --- | --- | --- | --- |
| Asian | 50 (5.2) | 85 (4.5) | 23 (27.1) | 10.6% | 9 | 2.6 (1.5-3.6) |
| Black | 15 (1.6) | 28 (1.5) | 8 (28.6) | 4.9% | 1 | 5.9 (1.8-9.9) |
| Mixed/other | 41 (4.2) | 76 (4.0) | 10 (13.2) | 13.6% | 10 | 1 (0.4-1.6) |
| White | 862 (90.0) | 1690 (89.9) | 1188 (70.3) | 71.0% | 1200 | 1 (0.9-1.0) |
| Total | 968 | 1879 | 1229 (65.4) |  |  |  |

\* n=50 (2.7%) of partners were reported to be of unknown ethnicity
\*\* Number of partners expected of the same ethnicity if there was no positive or negative tendency towards same ethnicity partnerships.
Ethnicity of participants is self-reported; ethnicity of partners is reported by participants.
Assumes that the population of potential male sexual partners has ethnicity distribution equal to that of RiSH reported partners.
Note that the overall 2021 ONS census population ethnicity distribution for England and Wales is 9.3% Asian, 4.0% Black, 2.9% Mixed, 81.7% White, 2.1% Other. If these figures are used to calculate homophily ratios we find: Asian 2.9 (1.7-4.1); Black 7.2 (2.2-12.1); Mixed/Other 2.6 (1.0-4.3) and White 0.9 (0.8-0.9).

## Discussion

This study has addressed a key evidence gap in our quantitative understanding of sexual partners among UK GBMSM. Previous research has highlighted the utility for sexual health in differentiating between different sexual partnership types, but this research was not specifically conducted among GBMSM(17, 18). Using data from a large community sample, a variety of sexual partnership types, distinguishable in lengths, overlaps, homophily, sexual behaviour and sexual health communication were reported. This heterogeneity underscores the importance of accounting for different types of partners in tailoring sexual health interventions (17) and in research to understand population effectiveness of sexual health interventions.

Some STI prevention strategies are negotiated between partners (e.g. condoms), some decided upon by individuals (e.g. HIV or STI prophylaxis) and some developed within relationships (e.g. couples negotiating openness and sexual health strategies). For over half of partners reported, sexual health communication was reported to be ‘easy’, but among one-time partnerships, two thirds had not involved sexual health communication. Low condom use within partnerships involving anal sex supports the need for new acceptable STI prevention strategies to supplement condoms that account for different communication patterns across partnership types.

Consistent with other studies, we found a highly skewed distribution of number of male sexual partners reported over the previous 3 months(6). We also found that participants had on average more than one ongoing sexual partnership at time of survey completion, and overlaps among recent sexual partners, indicators of partnership concurrency. These characteristics have in models been shown to facilitate rapid spread of epidemics(6, 19), and in some cases, raise endemic prevalence(20). These points should be appreciated in epidemic preparedness, particularly given that we also found substantial geographical mixing in partnerships.

Partnerships reported by GBMSM involved large mean but variable age differences, particularly among non-established partners. Similar mean age differences were also reported of most recent sexual partners among GBMSM in the United States (21). Age mixing could hypothetically affect STI transmission in the population either via differences in infectious prevalence by age, or via influences on STI prevention and sexual behaviour within a partnership(21). Further work would be needed to explore these questions among GBMSM and for different types of STIs in the UK; evidence about partnership age differences and sexual behaviour among GBMSM from other places is mixed ((22–24) (25)).

Our findings suggest that some ethnically minoritised GBMSM in the UK could be more likely to form partnerships with each other than would be expected based on overall population numbers. Our findings should be interpreted with caution given small numbers of participants from minoritized ethnicities, and participant reporting of partner ethnicities, though similar results have been observed in the United States (26). We have not explored the mechanisms behind ethnicity homophily, which could include racism and discrimination on dating apps(27), and/or seeking of others with similar life experiences. Ethnicity homophily among minoritized groups could potentially compound over the sexual network the effects of systematic inequities in sexual healthcare knowledge and access(28) and should be explored further.

Strengths of our study include co-design of the partnership module, embedded within an established community survey reaching beyond clinic-attending populations. However, recruitment was not population-representative and half of participants were recruited via Grindr, which could have biased our findings towards GBMSM with multiple recent partnerships. While the majority of eligible participants did complete optional 2^nd^ and 3^rd^ most recent partner questions, we cannot compare to the characteristics of partners not reported. The 2023 RiiSH survey had a similar demographic make-up to previous surveys(16), with high proportions of White, highly educated, and England-residing participants. Participants were on average in mid-life, though there is evidence that GBMSM partner numbers and concurrency peak around age 35-54, rather than declining from 30(29), as seen in heterosexuals .The inclusion of partnerships with younger men in our data also gives insight into this population.

## Conclusion

We have found that sexual partnership types among GBMSM are associated with different sexual behaviours, communication, use of STI prevention and relative partner demographics. These data can be used to inform our understanding of the contexts in which transmission and decision-making about STI prevention take place. Our findings bring an updated and more detailed understanding of GBMSM sexual partnership network characteristics and the interaction between network structure, demography and behaviour, critical to informing effective and equitable epidemic preparedness and combination STI prevention strategies.

## Data Availability

The data that support the findings of this study have been assessed by the UK Health Security Agency's Office for Data Acquisition and Release as having sensitive personal information and are therefore not publicly available to protect participant privacy. However, some aggregate data may be available upon reasonable request from the UKHSA. Requests can be directed to (UKHSA REGG Ref 524).

## Acknowledgements

We thank participants in the RiiSH 2023 survey for their time and sharing their personal information. We thank community members and organisations who contributed to the development of the survey module in 2023 and The Love Tank for supporting this process. The research was funded by the National Institute for Health Research Health Protection Research Unit (NIHR HPRU) in Blood Borne and Sexually Transmitted Infections at University College London in partnership with UKHSA, and by the Medical Research Council (MR/S020462/2). The views expressed are those of the authors and not necessarily those of the NIHR, the Department of Health and Social Care or UKHSA. We acknowledge members of the NIHR HPRU BBSTI Steering Committee: Professor Caroline Sabin (HPRU Director), Dr John Saunders (UKHSA Lead), Professor Catherine Mercer, Professor Gwenda Hughes, Dr Hamish Mohammed, Professor Greta Rait, Dr Ruth Simmons, Professor William Rosenberg, Dr Tamyo Mbisa, Professor Rosalind Raine, Dr Sema Mandal, Dr Rosamund Yu, Dr Samreen Ijaz, Dr Fabiana Lorencatto, Dr Rachel Hunter, Dr Kirsty Foster and Dr Mamooma Tahir.

## Appendix

**Figure A1:**
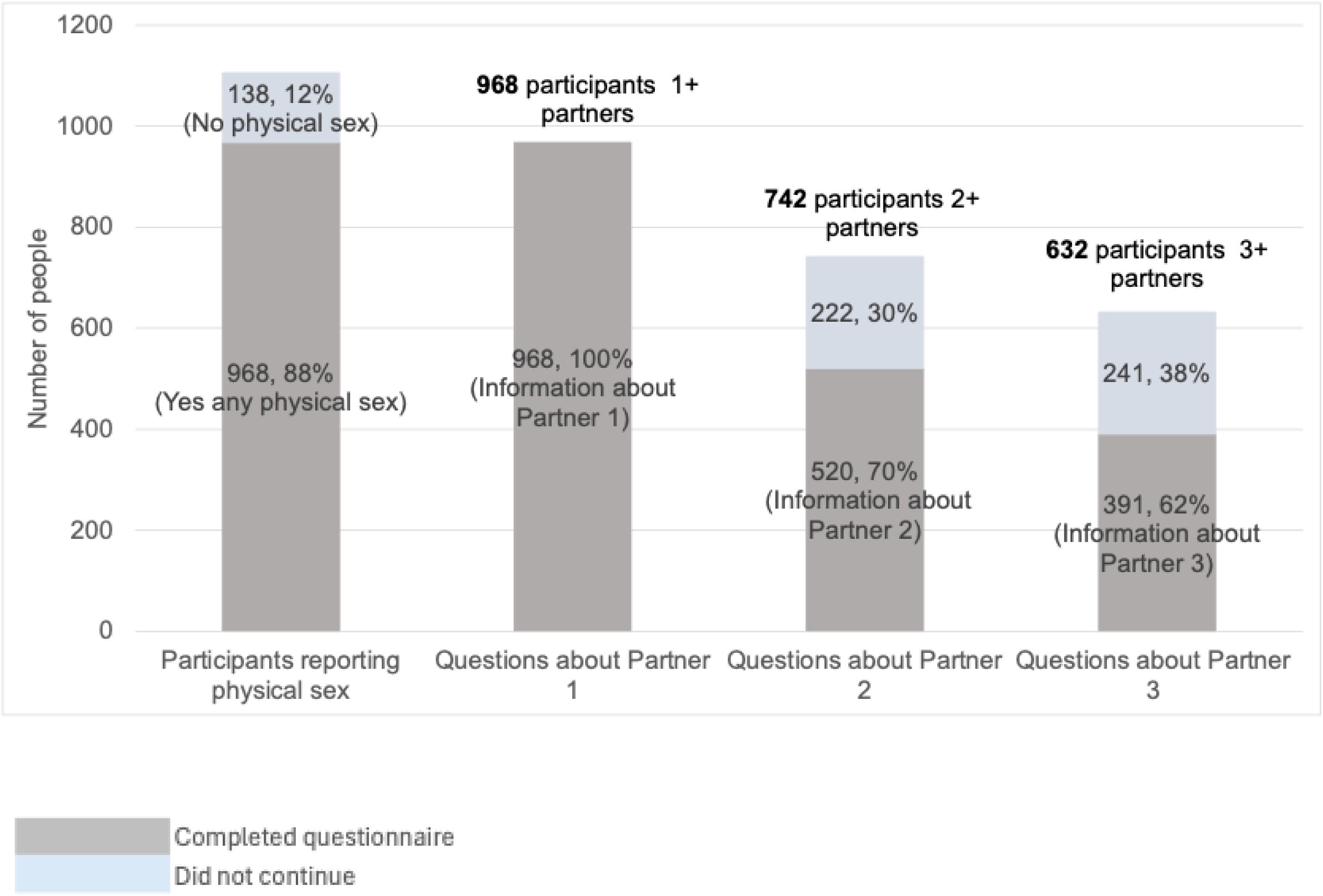
Most recent partners reported by RiiSH 2023 participants among those eligible

**Figure A2:**
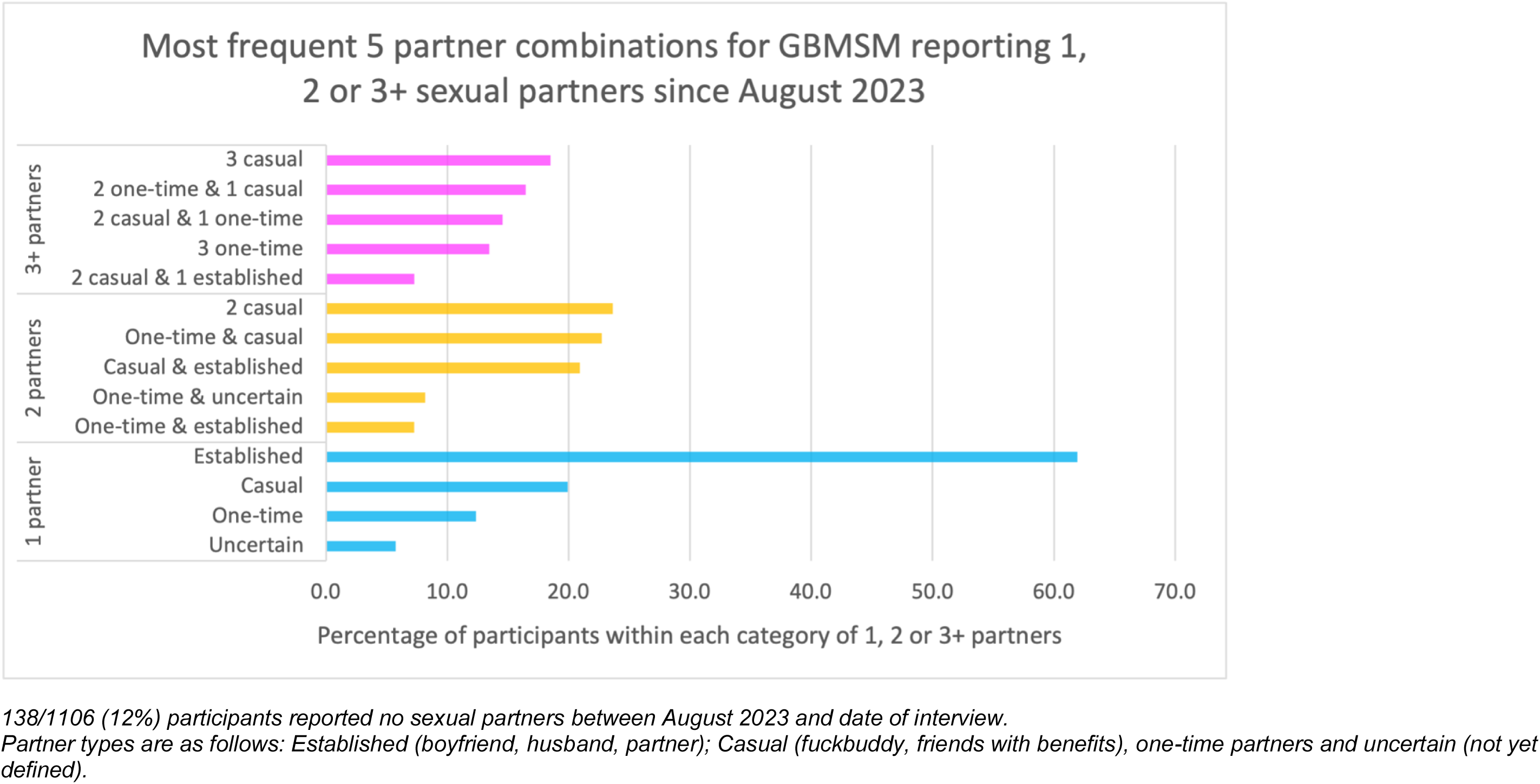
Frequent combinations of most recent sexual partner type among 968 GBMSM reporting 1, 2 or 3+ total sexual partners in the previous 3 months

**Figure A3:**
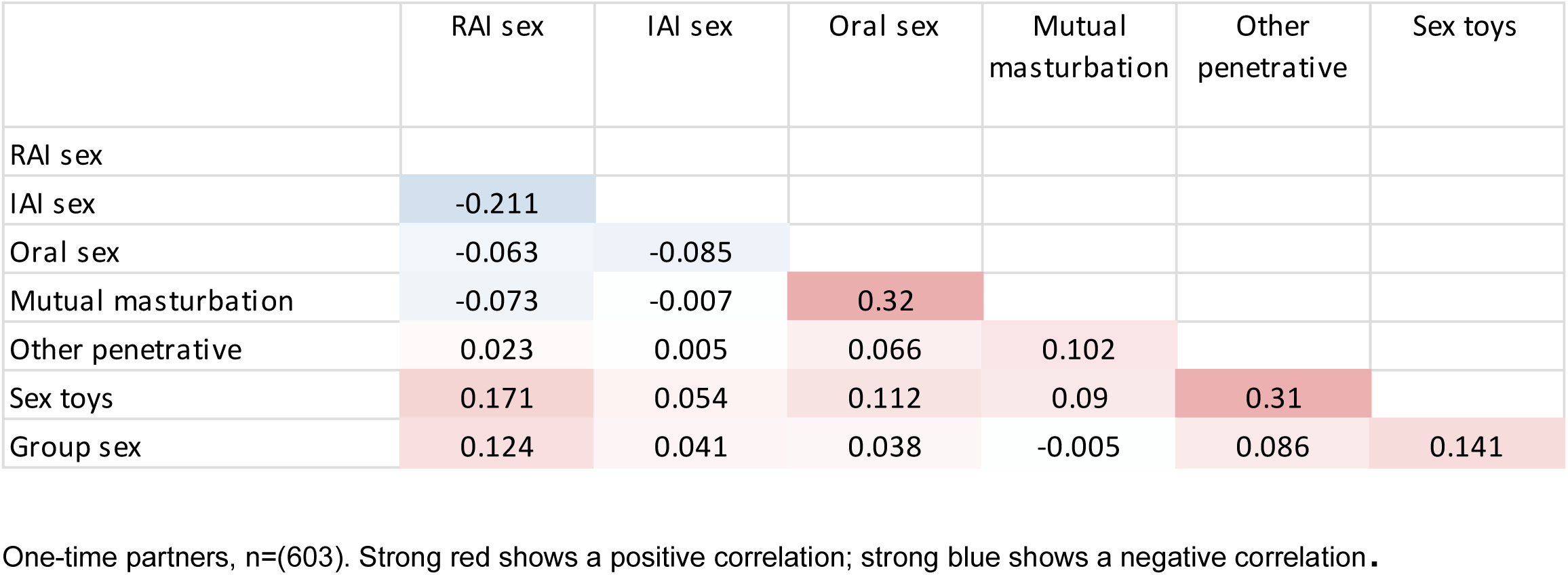
Pearson correlation coefficients between sexual behaviours with one-time sexual partners

**Figure A4:**
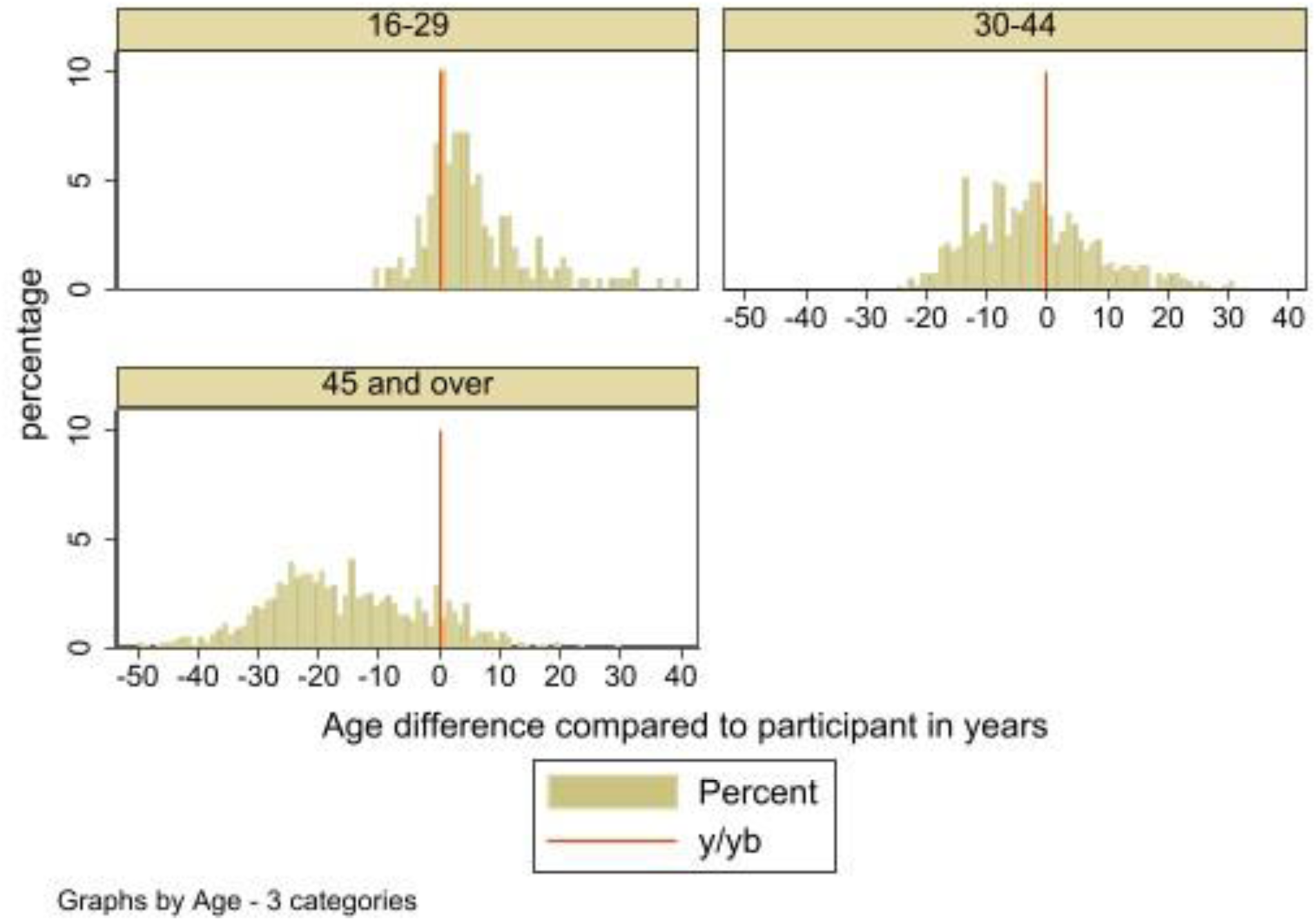
Partnership age gap distributions in years by participant age

**Table A1:** Numbers of sexual partners reported since August 2023 among all RiiSH participants.

| Number of physical male sexual partners since Aug 2023 (3 months) |  |  | Number of CAS male sexual partners since Aug 2023 |  |  | Number of physical female sexual partners since Aug 2023 |  |  |
| --- | --- | --- | --- | --- | --- | --- | --- | --- |
|  | n | % |  | n | % |  | n | % |
| 0 | 138 | 12.5 | 0 | 383 | 34.6 | 0 | 1016 | 91.9 |
| 1 | 226 | 20.4 | 1 | 270 | 24.4 | 1 | 61 | 5.5 |
| 2 | 110 | 10 | 2 | 102 | 9.2 | 2 | 14 | 1.3 |
| 3 | 102 | 9.2 | 3 | 71 | 6.4 | 3 | 5 | 0.5 |
| 4 | 65 | 5.9 | 4 | 49 | 4.4 | 4 | 1 | 0.1 |
| 5 | 77 | 7 | 5 | 39 | 3.5 | 5 | 1 | 0.1 |
| 6 | 58 | 5.2 | 6 | 31 | 2.8 | 6 | 0 | 0 |
| 7 | 22 | 2 | 7 | 19 | 1.7 | 7 | 1 | 0.1 |
| 8 | 30 | 2.7 | 8 | 11 | 1 | 8 | 0 | 0 |
| 9 | 8 | 0.7 | 9 | 9 | 0.8 | 9 | 0 | 0 |
| 10 | 53 | 4.8 | 10 | 20 | 1.8 | 10 | 0 | 0 |
| 11-20 | 111 | 10 | 11-20 | 52 | 4.7 | 11-20 | 4 | 0.4 |
| 21-30 | 48 | 4.3 | 21-30 | 24 | 2.2 | 21-30 | 1 | 0.1 |
| 31-40 | 20 | 1.8 | 31-40 | 8 | 0.7 | 31-40 | 0 | 0 |
| 41-50 | 10 | 0.9 | 41-50 | 9 | 0.8 | 41-50 | 0 | 0 |
| 50+ | 28 | 2.5 | 50+ | 9 | 0.8 | 50+ | 2 | 0.2 |

**Total physical male sexual partners reported among participants since August 2023:**
8793 (mid-point)
7400 (low estimate, taking lower bound of categorical responses)
9801 (higher estimate, taking higher bound of categorical responses)

**Table A2:**
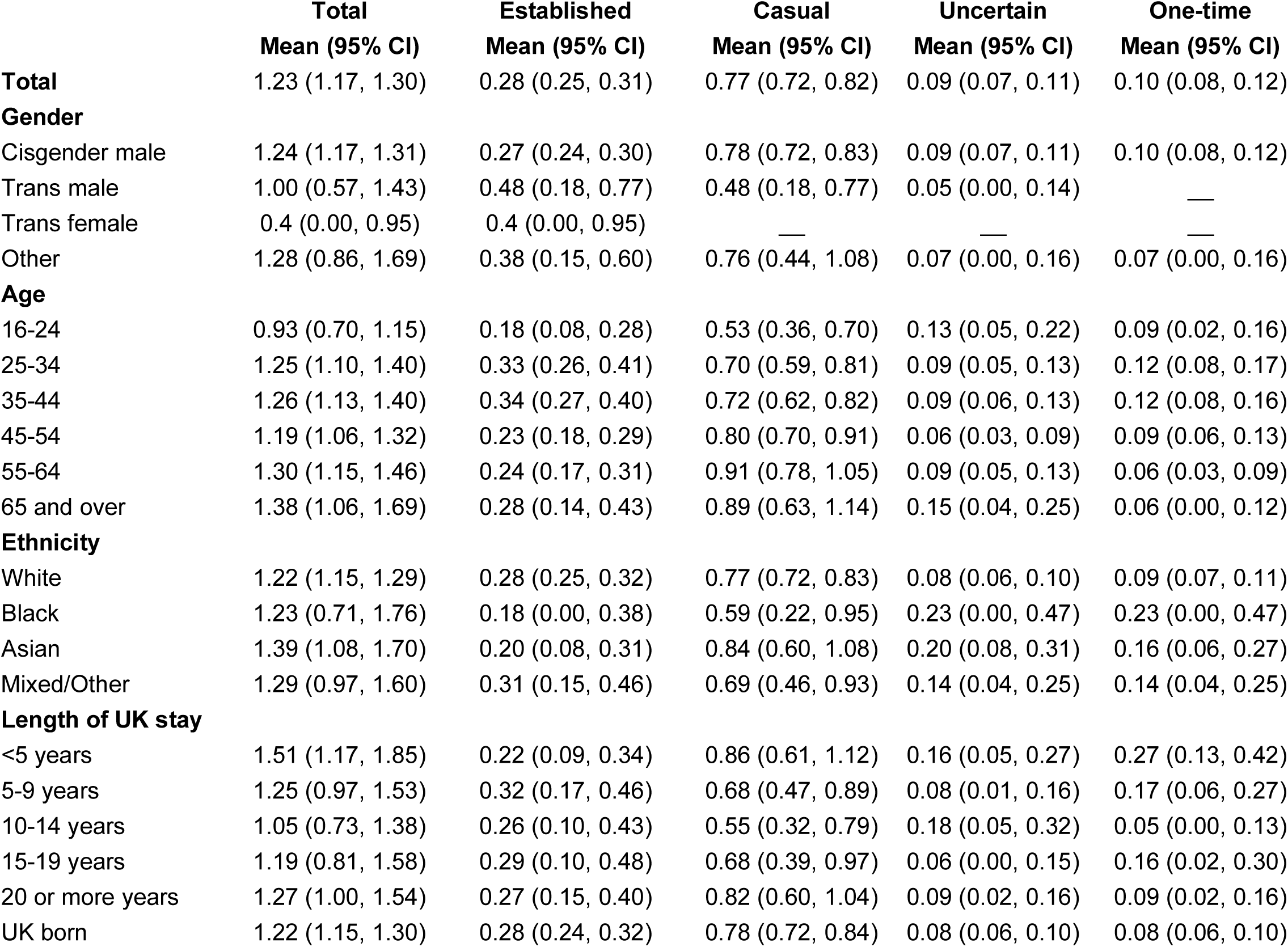
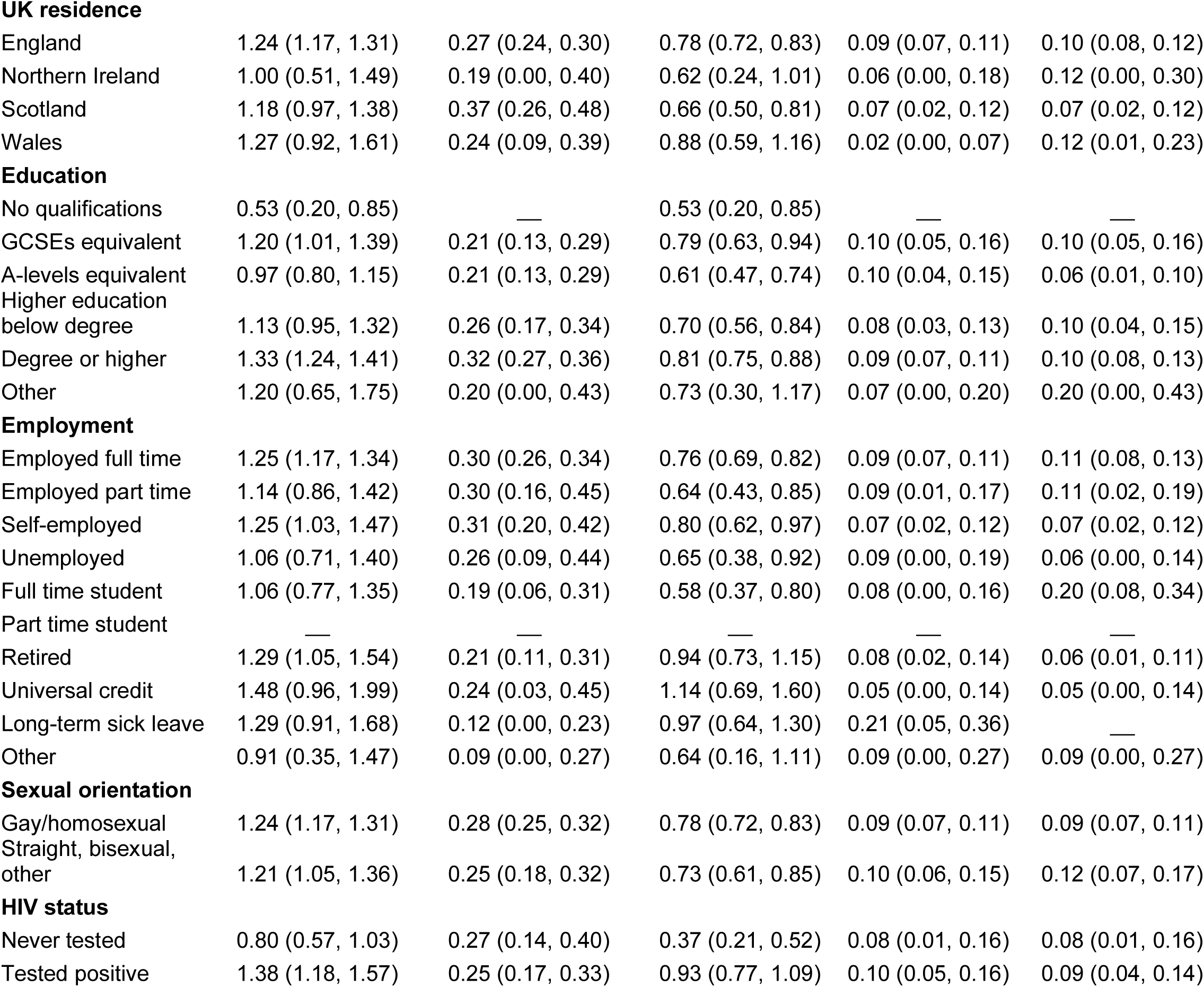
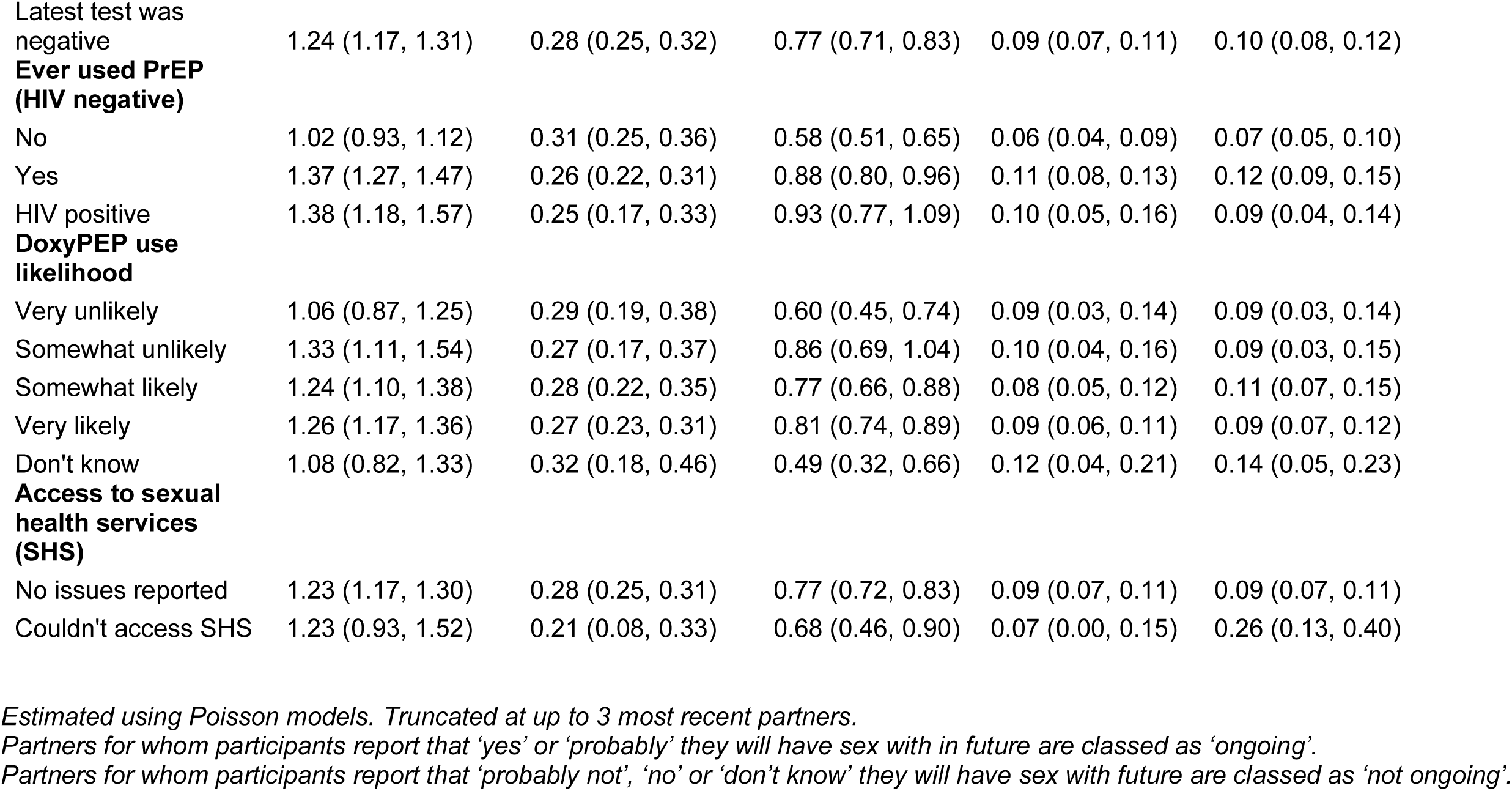
Mean momentary degree by participant characteristics (n=1106)

|  | **Total** | **Established** | **Casual** | **Uncertain** | **One-time** |
| --- | --- | --- | --- | --- | --- |
|  | **Mean (95% CI)** | **Mean (95% CI)** | **Mean (95% CI)** | **Mean (95% CI)** | **Mean (95% CI)** |
| **Total** | 1.23 (1.17, 1.30) | 0.28 (0.25, 0.31) | 0.77 (0.72, 0.82) | 0.09 (0.07, 0.11) | 0.10 (0.08, 0.12) |
| **Gender** |  |  |  |  |  |
| Cisgender male | 1.24 (1.17, 1.31) | 0.27 (0.24, 0.30) | 0.78 (0.72, 0.83) | 0.09 (0.07, 0.11) | 0.10 (0.08, 0.12) |
| Trans male | 1.00 (0.57, 1.43) | 0.48 (0.18, 0.77) | 0.48 (0.18, 0.77) | 0.05 (0.00, 0.14) | — |
| Trans female | 0.4 (0.00, 0.95) | 0.4 (0.00, 0.95) | — | — | — |
| Other | 1.28 (0.86, 1.69) | 0.38 (0.15, 0.60) | 0.76 (0.44, 1.08) | 0.07 (0.00, 0.16) | 0.07 (0.00, 0.16) |
| **Age** |  |  |  |  |  |
| 16-24 | 0.93 (0.70, 1.15) | 0.18 (0.08, 0.28) | 0.53 (0.36, 0.70) | 0.13 (0.05, 0.22) | 0.09 (0.02, 0.16) |
| 25-34 | 1.25 (1.10, 1.40) | 0.33 (0.26, 0.41) | 0.70 (0.59, 0.81) | 0.09 (0.05, 0.13) | 0.12 (0.08, 0.17) |
| 35-44 | 1.26 (1.13, 1.40) | 0.34 (0.27, 0.40) | 0.72 (0.62, 0.82) | 0.09 (0.06, 0.13) | 0.12 (0.08, 0.16) |
| 45-54 | 1.19 (1.06, 1.32) | 0.23 (0.18, 0.29) | 0.80 (0.70, 0.91) | 0.06 (0.03, 0.09) | 0.09 (0.06, 0.13) |
| 55-64 | 1.30 (1.15, 1.46) | 0.24 (0.17, 0.31) | 0.91 (0.78, 1.05) | 0.09 (0.05, 0.13) | 0.06 (0.03, 0.09) |
| 65 and over | 1.38 (1.06, 1.69) | 0.28 (0.14, 0.43) | 0.89 (0.63, 1.14) | 0.15 (0.04, 0.25) | 0.06 (0.00, 0.12) |
| **Ethnicity** |  |  |  |  |  |
| White | 1.22 (1.15, 1.29) | 0.28 (0.25, 0.32) | 0.77 (0.72, 0.83) | 0.08 (0.06, 0.10) | 0.09 (0.07, 0.11) |
| Black | 1.23 (0.71, 1.76) | 0.18 (0.00, 0.38) | 0.59 (0.22, 0.95) | 0.23 (0.00, 0.47) | 0.23 (0.00, 0.47) |
| Asian | 1.39 (1.08, 1.70) | 0.20 (0.08, 0.31) | 0.84 (0.60, 1.08) | 0.20 (0.08, 0.31) | 0.16 (0.06, 0.27) |
| Mixed/Other | 1.29 (0.97, 1.60) | 0.31 (0.15, 0.46) | 0.69 (0.46, 0.93) | 0.14 (0.04, 0.25) | 0.14 (0.04, 0.25) |
| **Length of UK stay** |  |  |  |  |  |
| <5 years | 1.51 (1.17, 1.85) | 0.22 (0.09, 0.34) | 0.86 (0.61, 1.12) | 0.16 (0.05, 0.27) | 0.27 (0.13, 0.42) |
| 5-9 years | 1.25 (0.97, 1.53) | 0.32 (0.17, 0.46) | 0.68 (0.47, 0.89) | 0.08 (0.01, 0.16) | 0.17 (0.06, 0.27) |
| 10-14 years | 1.05 (0.73, 1.38) | 0.26 (0.10, 0.43) | 0.55 (0.32, 0.79) | 0.18 (0.05, 0.32) | 0.05 (0.00, 0.13) |
| 15-19 years | 1.19 (0.81, 1.58) | 0.29 (0.10, 0.48) | 0.68 (0.39, 0.97) | 0.06 (0.00, 0.15) | 0.16 (0.02, 0.30) |
| 20 or more years | 1.27 (1.00, 1.54) | 0.27 (0.15, 0.40) | 0.82 (0.60, 1.04) | 0.09 (0.02, 0.16) | 0.09 (0.02, 0.16) |
| UK born | 1.22 (1.15, 1.30) | 0.28 (0.24, 0.32) | 0.78 (0.72, 0.84) | 0.08 (0.06, 0.10) | 0.08 (0.06, 0.10) |
| **UK residence** |  |  |  |  |  |
| England | 1.24 (1.17, 1.31) | 0.27 (0.24, 0.30) | 0.78 (0.72, 0.83) | 0.09 (0.07, 0.11) | 0.10 (0.08, 0.12) |
| Northern Ireland | 1.00 (0.51, 1.49) | 0.19 (0.00, 0.40) | 0.62 (0.24, 1.01) | 0.06 (0.00, 0.18) | 0.12 (0.00, 0.30) |
| Scotland | 1.18 (0.97, 1.38) | 0.37 (0.26, 0.48) | 0.66 (0.50, 0.81) | 0.07 (0.02, 0.12) | 0.07 (0.02, 0.12) |
| Wales | 1.27 (0.92, 1.61) | 0.24 (0.09, 0.39) | 0.88 (0.59, 1.16) | 0.02 (0.00, 0.07) | 0.12 (0.01, 0.23) |
| **Education** |  |  |  |  |  |
| No qualifications | 0.53 (0.20, 0.85) | — | 0.53 (0.20, 0.85) | — | — |
| GCSEs equivalent | 1.20 (1.01, 1.39) | 0.21 (0.13, 0.29) | 0.79 (0.63, 0.94) | 0.10 (0.05, 0.16) | 0.10 (0.05, 0.16) |
| A-levels equivalent | 0.97 (0.80, 1.15) | 0.21 (0.13, 0.29) | 0.61 (0.47, 0.74) | 0.10 (0.04, 0.15) | 0.06 (0.01, 0.10) |
| Higher education below degree | 1.13 (0.95, 1.32) | 0.26 (0.17, 0.34) | 0.70 (0.56, 0.84) | 0.08 (0.03, 0.13) | 0.10 (0.04, 0.15) |
| Degree or higher | 1.33 (1.24, 1.41) | 0.32 (0.27, 0.36) | 0.81 (0.75, 0.88) | 0.09 (0.07, 0.11) | 0.10 (0.08, 0.13) |
| Other | 1.20 (0.65, 1.75) | 0.20 (0.00, 0.43) | 0.73 (0.30, 1.17) | 0.07 (0.00, 0.20) | 0.20 (0.00, 0.43) |
| **Employment** |  |  |  |  |  |
| Employed full time | 1.25 (1.17, 1.34) | 0.30 (0.26, 0.34) | 0.76 (0.69, 0.82) | 0.09 (0.07, 0.11) | 0.11 (0.08, 0.13) |
| Employed part time | 1.14 (0.86, 1.42) | 0.30 (0.16, 0.45) | 0.64 (0.43, 0.85) | 0.09 (0.01, 0.17) | 0.11 (0.02, 0.19) |
| Self-employed | 1.25 (1.03, 1.47) | 0.31 (0.20, 0.42) | 0.80 (0.62, 0.97) | 0.07 (0.02, 0.12) | 0.07 (0.02, 0.12) |
| Unemployed | 1.06 (0.71, 1.40) | 0.26 (0.09, 0.44) | 0.65 (0.38, 0.92) | 0.09 (0.00, 0.19) | 0.06 (0.00, 0.14) |
| Full time student | 1.06 (0.77, 1.35) | 0.19 (0.06, 0.31) | 0.58 (0.37, 0.80) | 0.08 (0.00, 0.16) | 0.20 (0.08, 0.34) |
| Part time student | — | — | — | — | — |
| Retired | 1.29 (1.05, 1.54) | 0.21 (0.11, 0.31) | 0.94 (0.73, 1.15) | 0.08 (0.02, 0.14) | 0.06 (0.01, 0.11) |
| Universal credit | 1.48 (0.96, 1.99) | 0.24 (0.03, 0.45) | 1.14 (0.69, 1.60) | 0.05 (0.00, 0.14) | 0.05 (0.00, 0.14) |
| Long-term sick leave | 1.29 (0.91, 1.68) | 0.12 (0.00, 0.23) | 0.97 (0.64, 1.30) | 0.21 (0.05, 0.36) | — |
| Other | 0.91 (0.35, 1.47) | 0.09 (0.00, 0.27) | 0.64 (0.16, 1.11) | 0.09 (0.00, 0.27) | 0.09 (0.00, 0.27) |
| **Sexual orientation** |  |  |  |  |  |
| Gay/homosexual | 1.24 (1.17, 1.31) | 0.28 (0.25, 0.32) | 0.78 (0.72, 0.83) | 0.09 (0.07, 0.11) | 0.09 (0.07, 0.11) |
| Straight, bisexual, other | 1.21 (1.05, 1.36) | 0.25 (0.18, 0.32) | 0.73 (0.61, 0.85) | 0.10 (0.06, 0.15) | 0.12 (0.07, 0.17) |
| **HIV status** |  |  |  |  |  |
| Never tested | 0.80 (0.57, 1.03) | 0.27 (0.14, 0.40) | 0.37 (0.21, 0.52) | 0.08 (0.01, 0.16) | 0.08 (0.01, 0.16) |
| Tested positive | 1.38 (1.18, 1.57) | 0.25 (0.17, 0.33) | 0.93 (0.77, 1.09) | 0.10 (0.05, 0.16) | 0.09 (0.04, 0.14) |
| Latest test was negative | 1.24 (1.17, 1.31) | 0.28 (0.25, 0.32) | 0.77 (0.71, 0.83) | 0.09 (0.07, 0.11) | 0.10 (0.08, 0.12) |
| **Ever used PrEP (HIV negative)** |  |  |  |  |  |
| No | 1.02 (0.93, 1.12) | 0.31 (0.25, 0.36) | 0.58 (0.51, 0.65) | 0.06 (0.04, 0.09) | 0.07 (0.05, 0.10) |
| Yes | 1.37 (1.27, 1.47) | 0.26 (0.22, 0.31) | 0.88 (0.80, 0.96) | 0.11 (0.08, 0.13) | 0.12 (0.09, 0.15) |
| HIV positive | 1.38 (1.18, 1.57) | 0.25 (0.17, 0.33) | 0.93 (0.77, 1.09) | 0.10 (0.05, 0.16) | 0.09 (0.04, 0.14) |
| **DoxyPEP use likelihood** |  |  |  |  |  |
| Very unlikely | 1.06 (0.87, 1.25) | 0.29 (0.19, 0.38) | 0.60 (0.45, 0.74) | 0.09 (0.03, 0.14) | 0.09 (0.03, 0.14) |
| Somewhat unlikely | 1.33 (1.11, 1.54) | 0.27 (0.17, 0.37) | 0.86 (0.69, 1.04) | 0.10 (0.04, 0.16) | 0.09 (0.03, 0.15) |
| Somewhat likely | 1.24 (1.10, 1.38) | 0.28 (0.22, 0.35) | 0.77 (0.66, 0.88) | 0.08 (0.05, 0.12) | 0.11 (0.07, 0.15) |
| Very likely | 1.26 (1.17, 1.36) | 0.27 (0.23, 0.31) | 0.81 (0.74, 0.89) | 0.09 (0.06, 0.11) | 0.09 (0.07, 0.12) |
| Don't know | 1.08 (0.82, 1.33) | 0.32 (0.18, 0.46) | 0.49 (0.32, 0.66) | 0.12 (0.04, 0.21) | 0.14 (0.05, 0.23) |
| **Access to sexual health services (SHS)** |  |  |  |  |  |
| No issues reported | 1.23 (1.17, 1.30) | 0.28 (0.25, 0.31) | 0.77 (0.72, 0.83) | 0.09 (0.07, 0.11) | 0.09 (0.07, 0.11) |
| Couldn't access SHS | 1.23 (0.93, 1.52) | 0.21 (0.08, 0.33) | 0.68 (0.46, 0.90) | 0.07 (0.00, 0.15) | 0.26 (0.13, 0.40) |

*Estimated using Poisson models. Truncated at up to 3 most recent partners.*
*Partners for whom participants report that 'yes' or 'probably' they will have sex with in future are classed as 'ongoing'.*
*Partners for whom participants report that 'probably not', 'no' or 'don't know' they will have sex with future are classed as 'not ongoing'.*

